# Deep Learning Model for Mortality Prediction of ICU Patients with Paralytic Ileus

**DOI:** 10.1101/2021.10.06.21264665

**Authors:** Martha Razo, Maryam Pishgar, Houshang Darabi

## Abstract

**Background:** Paralytic Ileus (PI) patients in the Intensive Care Unit (ICU) are at a significant risk of death. Prediction of at-risk patients for mortality after 24 hours of admission of ICU PI patients is important to increase the life expectancy of PI patients.

**Methods and Results:** The proposed framework, *DLMP* (Deep Learning Model for Mortality Prediction of ICU Patients with PI) is a powerful deep learning model consisting of six total unique clinical lab items and two demographics as inputs to a Neural Network(NN) of only two neuron layers. Using the Medical Information Mart for Intensive Care III (MIMIC-III) dataset of 1,017 ICU PI patients, the *DLMP* resulted in the best prediction performance with an AUC score of 0.866.

**Conclusion:** The proposed approach is capable of modeling the mortality of ICU patients after 24 hours admission using only six unique total clinical data and two demographics with a simple NN architecture. *DLMP* framework significantly improves the outcome prediction compared to the process mining and machine learning models. The proposed *DLMP* has the potential of allowing clinicians to create targeted interventions that reduce mortality for PI patients in an ICU setting.

## 1 Introduction

PI is an intestinal obstruction, intestinal contents fail to progress, causing abdominal pain and vomiting [1]. PI patients are at risk of mortality as high as 45% in the ICU setting [2]. PI patients who are admitted to ICU are especially at risk of dying because of the seriousness of their condition [3]. Early prediction of the outcome of patients diagnosed with PI in the ICU could be helpful for an appropriate treatment plan for patients. It is key that current research focuses on creating accurate models for predicting mortality of ICU patients with PI to increase patients’ life span.

Several existing models have been developed to predict the mortality of ICU patients diagnosed with PI. However, these models use numerous variables and the architecture of the models are very complex. Moreover, the results of the existing models are not reliable enough for such a serious health condition as PI.

This work focuses on determining the mortality of ICU patients with PI after 24 hours of being admitted using a deep learning model, that uses as inputs only 6 unique total clinical lab items, and 2 demographic information. The number of variables is reduced to the most significant variables for predicting PI patient mortality. The proposed framework is called *DLMP* (Deep Learning Model for Mortality Prediction of ICU Patients with Paralytic Ileus) and it demonstrates significant performance for prediction.

## 2 Methodology

### 2.1 Data Source and Inclusion Criteria

Three distinct data sets were extracted from the MIMIC III datasets to create the *DLMP* framework. MIMIC III is a large database containing information relating to patients admitted to Beth Israel Deaconess Medical Center (BIDMC) [4]. A subset of 1,067 patients is selected using the ICD-9 code [5] from the MIMIC-III database. Furthermore, the PI patients under 18 years of age at their first admission, and who died before 24 hours of being admitted to the ICU were excluded to create a final dataset of 1,017 patients. The dataset was shared by Ahmad (2020) which include 17 lab items and 4 demographics [6].

#### Lab Items

The 17 lab items include: Creatinine, Hematocrit, Hemoglobin, Platelet, Ptt, Potassium, Aniongap, Bun, Bilirubin, Sodium, Pt, Bicarbonate, Albumin, Lactate, White Blood count, Inr, and Glucose.

#### Patient Demographics

The 4 demographic information of each patient consists of: Age, Insurance, Gender, and Ethnicity.

#### Event Frequencies

The sequence of observations on each patient is defined as event logs. Each of these observations is defined as an event. The event logs used originate from the work in [7] which contains 49 unique events. Out of 49 distinct events, 3 of them belong to the admission type, 10 of them are related to careunit activities, and represent the specific location in the ICU, patients came in and out after being admitted; CCU, CSRU, MICU, SICU, and TSICU or left the aforementioned places. Moreover, 34 of the distinct events belong to the lab measurements which 17 of them are flagged as normal and 17 of them are flagged as abnormal lab measurements. Finally, 2 of the events are the type of discharge, either death or discharge for each patient. The discharge events were excluded since these are the target events for prediction. As a result a total of 47 event’s frequencies were used that includes the following events: 17 lab items, Abnormal lab items(ABN) (17), care unit activities(10), and ICU type on admission (3). The frequency of each event was calculated by summing the total times an event occurred in the event logs.

### 2.2 Variable Importance

The number of variables was reduced using cox-regression analysis and Kaplan-Meier survival analysis based on [6]. Furthermore, univariate analysis was created using a threshold of p-value = 0.02 to identify the significant variables associated with the predictor. As a result, twelve total variables were selected: five total clinical abnormal lab items (Anion gap, Platelet, PTT, Blood urea nitrogen, and Bilirubin), five lab event frequencies (Bicarbonate, Bilirubin, Blood urea nitrogen, Platelets, and PTT), and two demographics (age and insurance). Furthermore, variable importance using Random Forest was used. As a result, the abnormal lab event of bicarbonate event was removed. Finally, the trade off analysis between the number of variables and AUC score was conducted to minimize the number of variables while maintaining a high AUC score. The resulting data consists of the 12 total aforementioned variables associated with each patient.

### 2.3 Prediction of Mortality for PI patients

We propose a deep learning model for predicting the mortality of patients diagnosed with PI after 24 hours of admission to the ICU. Furthermore, several machine learning models were developed and used as baseline models for comparison of performance. The 12 variables, five total clinical abnormal lab items, the five lab event frequencies and the two demographics (age and insurance) are fed into a NN to predict. Figure 1 illustrates the overview of the proposed model framework. The same dataset was used to build the baseline models.

### 2.4 Statistical Analysis between Cohorts

The training and validation cohorts are compared using Chi-Square for categorical variables and two-sided t-tests for continuous variables. The significant level is determined based on *p <* 0.01. Descriptive statistics, model development, and statistical analysis are conducted using Python, version 3.7.

### 2.5 Variables Impacts

SHAP (SHapley Additive exPlanations) assigns each variable an importance value for a particular prediction [8]. With the SHAP analysis we are able to interpret how each variable contributes to the prediction of mortality for ICU PI patients.

### 2.6 Ablation Analysis

The ablation analysis was conducted for each part of the NN architecture including; number of layers, the number of neurons, the learning rate, the activation function, epochs and batch size to optimize the prediction performance. Figure 2 illustrates the deep learning model architecture.

## 3 Results

### 3.1 Cohorts Details

The description of the training and validation cohorts is presented in Table 1. The selected cohort of 1,017 patients was randomly split into a training and testing split using a 67/33 ratio producing a train set of 681 patients and test set of 336 patients. Furthermore, the train set was randomly split using an 80/20 ratio to obtain a train and validation split. The validation set contained 136 patients.

**Table 1.** Comparison of Variables in Training & Validation cohorts

| Characteristics | Training Cohort<br>(N=681) | Validation Cohort<br>(N=136) | p-value |
| --- | --- | --- | --- |
| **Outcome Variable N (%)** |  |  |  |
| Mortality | 16.6 (113) | 12.5 (17) | < 0.05 |
| **Demographics** |  |  |  |
| Insurance N (%) |  |  | < 0.05 |
| Insurance class 0 | 2.93(20) | 2.20 (3) |  |
| Insurance class 1 | 9.83(67) | 9.55 (13) |  |
| Insurance class 2 | 50.8(346) | 50 (68) |  |
| Insurance class 3 | 35.5(242) | 38.2 (52) |  |
| Insurance class 4 | 0.88(6) | 0 (0) |  |
| Age Mean(Std) | 61.8 (15.5) | 61.5 (15.9) | 0.871 |
| **Laboratory Findings N (%)** |  |  |  |
| *aniongap* |  |  | < 0.05 |
| aniongap_Abnormal | 22.8 (155) | 17.8 (24) |  |
| aniongap_Normal | 74.4(507) | 78.7 (107) |  |
| aniongap_Missing | 2.79(19) | 3.68 (5) |  |
| *platelet* |  |  | < 0.05 |
| platelet_Abnormal | 56.2 (383) | 55.9 (76) |  |
| platelet_Normal | 41.7 (284) | 41.2(56) |  |
| platelet_Missing | 2.06(14) | 2.94 (4) |  |
| *ptt* |  |  | < 0.05 |
| ptt_Abnormal | 44.3 (302) | 43.4 (59) |  |
| ptt_Normal | 43.6 (297) | 41.2 (56) |  |
| ptt_Missing | 12.0 (82) | 15.4 (21) |  |
| *bun* |  |  | < 0.05 |
| bun_Abnormal | 52.3 (356) | 51.5 (70) |  |
| bun_Normal | 45.8 (312) | 45.6 (62) |  |
| bun_Missing | 1.91 (13) | 2.94 (4) |  |
| *bilirubin* |  |  | < 0.05 |
| bun_Abnormal | 40.4 (275) | 48.5 (66) |  |
| bun_Normal | 36.0 (245) | 28.7 (39) |  |
| bun_Missing | 23.6 (161) | 22.8 (3) |  |
| **Frequencies of Lab Items N (%)** |  |  | < 0.05 |
| bicarbonate (frequency=0) | 31.3 (213) | 30.9 (42) |  |
| bicarbonate (frequency=1) | 26.9 (183) | 30.1 (41) |  |
| bicarbonate (frequency=2) | 26.1 (178) | 25.7 (35) |  |
| bilirubin(frequency=0) | 56.7 (386) | 64.0 (87) |  |
| bilirubin (frequency=1) | 27.5 (187) | 22.1 (30) |  |
| bilirubin(frequency=2) | 12.8 (87) | 11.8 (16) |  |
| bun(frequency=0) | 41.0 (279) | 40.0 (53) |  |
| bun (frequency=2) | 16.2 (170) | 15.4 (40) |  |
| bun(frequency=1) | 25.0 (110) | 29.4 (21) |  |
| platelet(frequency=2) | 28.9 (200) | 27.9 (40) |  |
| platelet(frequency=0) | 20.1 (197) | 20.6 (38) |  |
| platelet(frequency=1) | 29.4 (137) | 29.4 (28) |  |
| ptt(frequency=0) | 40.1 (276) | 44.1 (40) |  |
| ptt(frequency=1) | 31.1 (212) | 31.2 (43) |  |
| ptt(frequency=2) | 17.3 (118) | 17.7 (24) |  |

The variables that were selected are the following: two demographics data (Age and Insurance), five total clinical abnormal lab items (Aniongap, Bilirubin, Bun, Platelet, Ptt), and five lab event frequencies (Bicarbonate, Bilirubin, Bun, Platelet, and PTT).

In terms of age, the validation cohort (61.8 years) is about the same as the training cohort (61.5 years) with a p-value of 0.87 which indicates that there are no significant differences between the cohorts. In terms of insurance, the p-value is less than 0.05 which shows that the entire distribution of the insurance is significantly different between the cohorts. The details for the cohorts for the insurance classes are shown in Table 1. Furthermore, the abnormal lab items significantly differ between the cohorts with a p-value less than 0.05, of which the details are shown in Table 1. The frequencies of lab events in the training and validation cohorts are also significantly different between cohorts. The p-values values for the frequencies of lab events are less than 0.05.

### 3.2 NN Architecture Ablation Analysis

This section discusses the iterative procedure for creating the architecture of the *DLMP* NN. The first step for developing the NN was tuning the layers and number of neurons. Second, finding the learning rate that produced the best model performance. Moreover, the activation function used for the deep learning. Lastly, the epochs and batch size selected is summarized.

#### 3.2.1 Layers & Neurons

The NN for the *DLMP* framework was initialized using the 12 input variables for the input layer, and two hidden layers of 100 and 56 neurons respectively. The output layer consisted of one neuron with 2 possible outputs, 1 indicates that a patient died and 0 that a patient is discharged and lives. The NN was run with an epoch of 50, batch size of 40, and learning rate of 1e-4 using Adam as the optimizer. Adding and removing layers from the two hidden layers did not improve the AUC score of the NN for predicting the mortality of ICU patients with PI after 24 hours of being admitted. Hence, only two layers were used for the NN final architecture. The results for changing the number of layers are shown in Table 2. Next, was to find the appropriate number of neurons that will increase the model performance. The different tests are shown in Table 3. The final number architecture for the NN was two hidden layers with 156 and 32 neurons respectively.

**Table 2.** Test 1: Number of Layers

| Number of Layers | AUC |
| --- | --- |
| (12, 100, 56, 1) | 0.905 |
| (12, 100, 56, 18, 1) | 0.766 |
| (12, 100, 56, 18, 6, 1) | 0.782 |
| (12, 100, 1) | 0.730 |
| (12, 56, 1) | 0.690 |

**Table 3.** Test 2: Number of Neurons

| Number of Neurons | AUC |
| --- | --- |
| (12, 100, 56, 1) | 0.790 |
| (12, 130, 56, 1) | 0.776 |
| (12, 150, 56, 1) | 0.769 |
| (12, 156, 80, 1) | 0.749 |
| (12, 156, 30, 1) | 0.783 |
| (12, 156, 32, 1) | 0.798 |
| (12, 80, 56, 1) | 0.753 |
| (12, 90, 56, 1) | 0.755 |

#### 3.2.2 Learning Rate

Increasing and decreasing the learning rate worsen the AUC score. The the chosen learning rate for the NN was 5e-4 using Adam as the optimizer. The tests for the learning rate are summarized in Table 4.

**Table 4.**
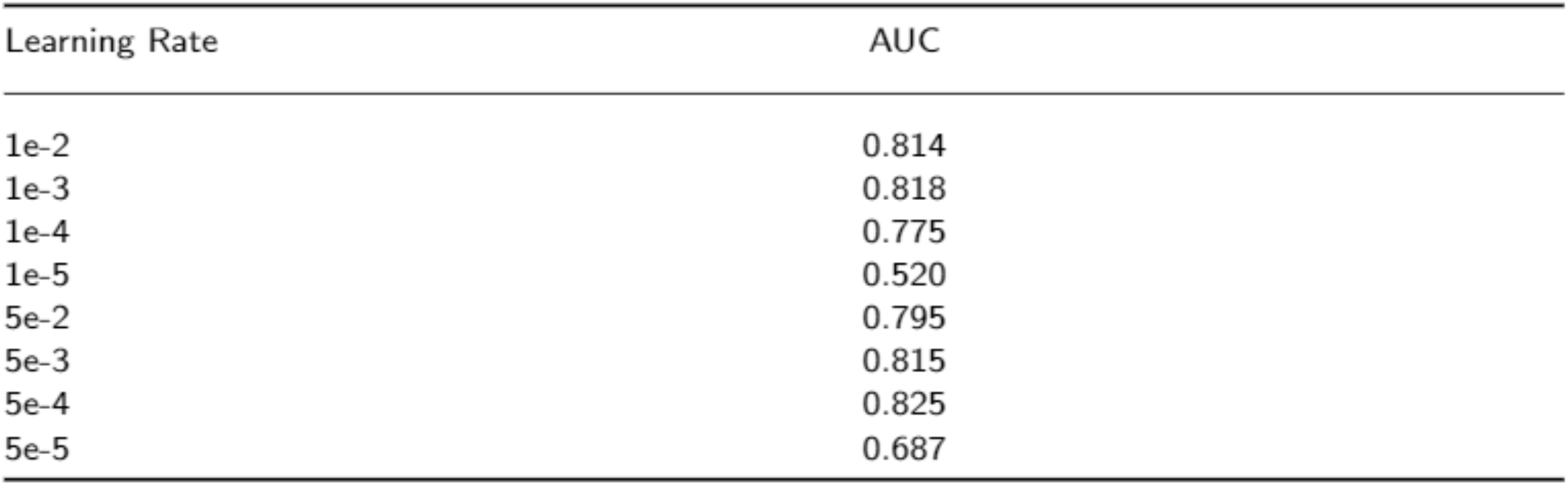
Test 3: Learning Rate

#### 3.2.3 Activation Function

Different combinations of activation functions were used, using the architecture (12,156, 32, 1) with learning rate of 5e-4. The activation functions used were Relu, Sigmoid and Tanh. All the following nine possible combinations were tested: Relu-Relu, Sigmoid-Relu, Tanh-Relu, Relu-Sigmoid, Relu-Tanh, Tanh-Sigmoid, Sigmoid-Tanh, Tanh-Tanh, and Sigmoid-Sigmoid were used for the two hidden layers and the best performance. Note since the output is categorical, discharged or dead, the most appropriate activation function for the output layer was Sigmoid. After running all nine possible combinations of activation functions for the two hidden layers, the Relu activation function for both hidden layers resulted in the best results. The test results are summarized in Table 5.

**Table 5.** Test 4: Activation Function Combinations

| Activation Function | AUC |
| --- | --- |
| Relu-Relu | 0.847 |
| Sigmoid-Relu | 0.817 |
| Tanh-Relu | 0.825 |
| Relu-Sigmoid | 0.837 |
| Relu-Tanh | 0.831 |
| Tanh-Sigmoid | 0.830 |
| Sigmoid-Tanh | 0.812 |
| Tanh-Tanh | 0.825 |
| Sigmoid-Sigmoid | 0.819 |

#### 3.2.4 Epochs & Batch Size

Finally, different epochs and bath size combinations were tested. using the architecture (12, 156, 32, 1) with learning rate of 5e-4 with Relu as the activation function for the two hidden layers. It was found that the best performance was achieved using an epoch of 150 and batch size of 30. The different tests that were run are listed in Table 6.

**Table 6.** Test 5: Epochs / Batch Size

| (Epochs, Batch Size) | AUC |
| --- | --- |
| (50, 40) | 0.841 |
| (50,30) | 0.859 |
| (50,20) | 0.857 |
| (100,50) | 0.858 |
| (100,40) | 0.852 |
| (100,30) | 0.874 |
| (150,30) | 0.910 |
| (150,50) | 0.896 |
| (200,30) | 0.868 |
| (200,50) | 0.882 |

The final NN for the *DLMP* framework architecture consisted of 2 hidden layers of 156 and 32 neurons respectively. The hidden layers use Re-lu as activation functions, and the output layer uses a sigmoid function. The NN used a learning rate of 5e-4 with Adam as the learning optimizer, epochs of 150, and batch size of 30. The deep learning model architecture for the *DLMP* is visualized in Figure 2.

### 3.3 SHAP Analysis

The results of SHAP analysis are shown in Figure 3. The correlations are colored, red means a feature is positively correlated with mortality and blue represents a negative correlation with the prediction.

Based on Figure 3. The variable’s contribution to the prediction are in the following order: Age, ppt Abnormal1, bilirubin Abnormal1, bun Abnormal1, bun, bilirubin, platelet, aniongap Abnormal1, ptt, insurance, bicarbonate, and platelet Abrnomal1.

Patients that have abnormal ptt, bilirubin, and anion gap clinical lab results have a high impact of death in the ICU setting. Moreover, an increased frequency of ptt lab tests can lead to a higher chance of death for a patient. The variables, bun Abrnormal1, bun, biliriub, and platelet are negatively correlated to the prediction. Furthermore, it is interpreted from the SHAP analysis that the least contributors for the prediction are bicarbonate frequency, a patient’s insurance, and having an abnormal platelet lab result. Lastly, having a normal bun lab result has a positive correlation to the prediction.

### 3.4 Evaluation Metrics and Proposed Model Performance

The proposed model is evaluated using the Area Under the Receiver Operating Characteristic curve (AUROC) [9] on the test cohort. AUROC metric calculates the rate of true positive over false positive for several threshold values. DeLong’s method is used to obtain 95% Confidence Intervals (CIs) of the AUROC value [10].

The *DLMP* frameowork is built into two phases. In Phase I the significant variables are determined. In Phase II, the optimal architecture for the NN for the *DLMP* is created. The *DLMP* framework has the best performance compared to other works and baseline machine models. The model proposed predicts with an AUC score of 86.6% and 95% CI of (0.817, 0.916). The results for the *DLMP* prediction framework are summarized in Table 7 and compared to the developed machine learning baseline models.

**Table 7.** *DLM P* & Machine Learning Baseline Models

| Prediction Model | AUC | AUC 95%*CI* |
| --- | --- | --- |
| *DLMP* | 0.866 | (0.817, 0.916) |
| Random Forest | 0.830 | (0.762, 0.890) |
| Gradient Boost. | 0.821 | (0.761, 0.880) |
| XGBoost | 0.820 | (0.755, 0.870) |
| Cat Boost. | 0.813 | (0.753, 0.872) |

## 4 Discussion

### 4.1 SHAP Analysis Interpretation Summary

From the SHAP mean score we can say that an older patient with PI after 24 hours of being admitted to the ICU has a higher chance of dying. We also note that having an abnormal ptt lab result should raise a flag for clinicians especially since ptt Abnormal1 has the highest contribution for prediction of death compared to having abnormal bilirubin, bicarbonate, and anion gap. If clinicians are limited in time and tests, then testing for ptt can be the first test to give a patient with PI. An increased frequency of ptt lab tests can possibly mean that there is unfortunately not much that can be done for the patient. Taking more ptt lab tests is just not going to be the answer for assuring the patient stays alive. Having fewer bun, bilirubin and platelet lab tests can mean that a patient will most likely die. For clinicians this means that increasing the frequency for the bun, bilirubin and platelet tests can help increase the probability of the patient to be discharged.

### 4.2 Existing Model Compilation Summary

There are two current existing papers that have been developed to predict the mortality of ICU patients diagnosed with PI that use MIMIC III dataset.

The first paper, developed the SRML-MortalityPredictor [6] framework with that used 17 variables, 14 abnormal clinical lab items and 3 demographics. Multiple machine learning models were developed to predict mortality. Support vector machine led to the best model performance with an AUC score of 81.38% [6].

The second work, PMPI (Process Mining Model to predict mortality of PI patients) uses DREAM (Decay Replay Mining) [10] and 49 total input variables consisting of patient medical history, the time related to the events, and demographic information for prediction [7]. PMPI resulted in similar if not better performance compared to the SRML-MortalityPredictor [6] with an AUC score of 0.82.

In this study, we investigated a deep learning model for predicting mortality of ICU patients with PI after 24 hours of admission, in which five abnormal lab items, five frequencies of lab events, and two demographics are fed into a NN model of two neuron layers. Also, several machine learning baseline models were developed for comparison.

The *DLMP* framework outperforms the best results of the existing literature in terms of the AUC score value proposed by [6] and [7]. The efficacy of our approach is demonstrated by a substantial improvement of at least +4.6% on the AUC score.

The existing proposed prediction models in the literature are able to predict mortality for ICU PI patients. However, these models have some disadvantages. First, the existing models use a multitude of variables for prediction. Ahmad 2020 uses a total of 17 variables, and Pishgar (2021) uses 49 variables for prediction. On the contrary, the proposed approach uses only 12 total variables which consist of only 6 total unique lab items information: Anion gap (abnormal lab event), Platelet (abnormal lab event and frequency), PTT (abnormal lab event and frequency), Blood urea nitrogen (abnormal lab event and frequency), Bilirubin (abnormal lab event and frequency), and Bicarbonate (frequency). This is crucial, since having fewer lab items for prediction has the potential of allowing clinicians to asses a patients mortality with less lab tests. This reduces the number of resources clinicians use to make decisions regarding treatment for patients. In affect, fewer input variables has the capacity of reducing costs for the hospital or medical institution while increasing the life expectancy of ICU PI patients.

Finally, the existing models are complex in their nature. For instance, process mining has multiple steps: process discovery, creating of timed state samples, and then these timed state samples are fed into a NN. The *DLMP* takes the data without modifications and uses only two neuron layers for prediction. The simplicity of the NN allows for quick results which can be generated using any machine and can be extracted in seconds. Every minute counts in decision making for patients to stay alive, clinicians cannot be waiting for extended periods of time for a model to give them prediction results.

### 4.3 Study Limitations

The proposed approach has one main limitation, our model uses the MIMIC III data set for model development and model evaluation. Using an independent dataset from a different hospital would be optimal to test the performance of the model [11], which provides room for future work.

## 5 Conclusion

PI patients are at high risk of death when admitted to the ICU if not treated immediately. This paper demonstrates significant performance improvements in predicting the mortality of ICU patients diagnosed with PI after 24 hours of being admitted. The proposed framework, *DLMP* prediction uses two phases for prediction. In Phase I the significant variables for prediction are determined. In Phase II, the optimal parameters for the deep learning NN are discovered iteratively changing the NN parameters. *DLMP* framework predicts mortality the best with an AUC score of 0.866.

The deep learning model in the *DLMP* framework was fed with only; 5 abnormal lab items, 2 demographic, and 5 lab event frequencies. This is a powerful generalized robust model. This means the model is fast performing and less complex than current proposed models for predicting mortality for ICU patients with PI. With the proposed *DLMP* prediction framework, doctors will use fewer labs to predict the mortality of patients. Less variables implies less time spent, fewer resources, less spending, and best more accurate decision making on patients with PI mortality. Future work will be to use another hospital’s data set for testing the proposed model. Further work will be to examine other modeling methods. Also, future work will be to further examine the six different lab items (Bicarbonate, Bilirubin, Bun, Platelet, Ptt, and Aniongap)in relation to predicting mortality for ICU patients with PI. It will be of value to partner with a medical expert to evaluate these six lab items in respect to PI diagnosis and mortality prediction.

## Data Availability

MIMIC III dataset is a publically available data

## Author details

University of Illinois at Chicago, Pishgar and Razo had equal contribution to this paper.

---

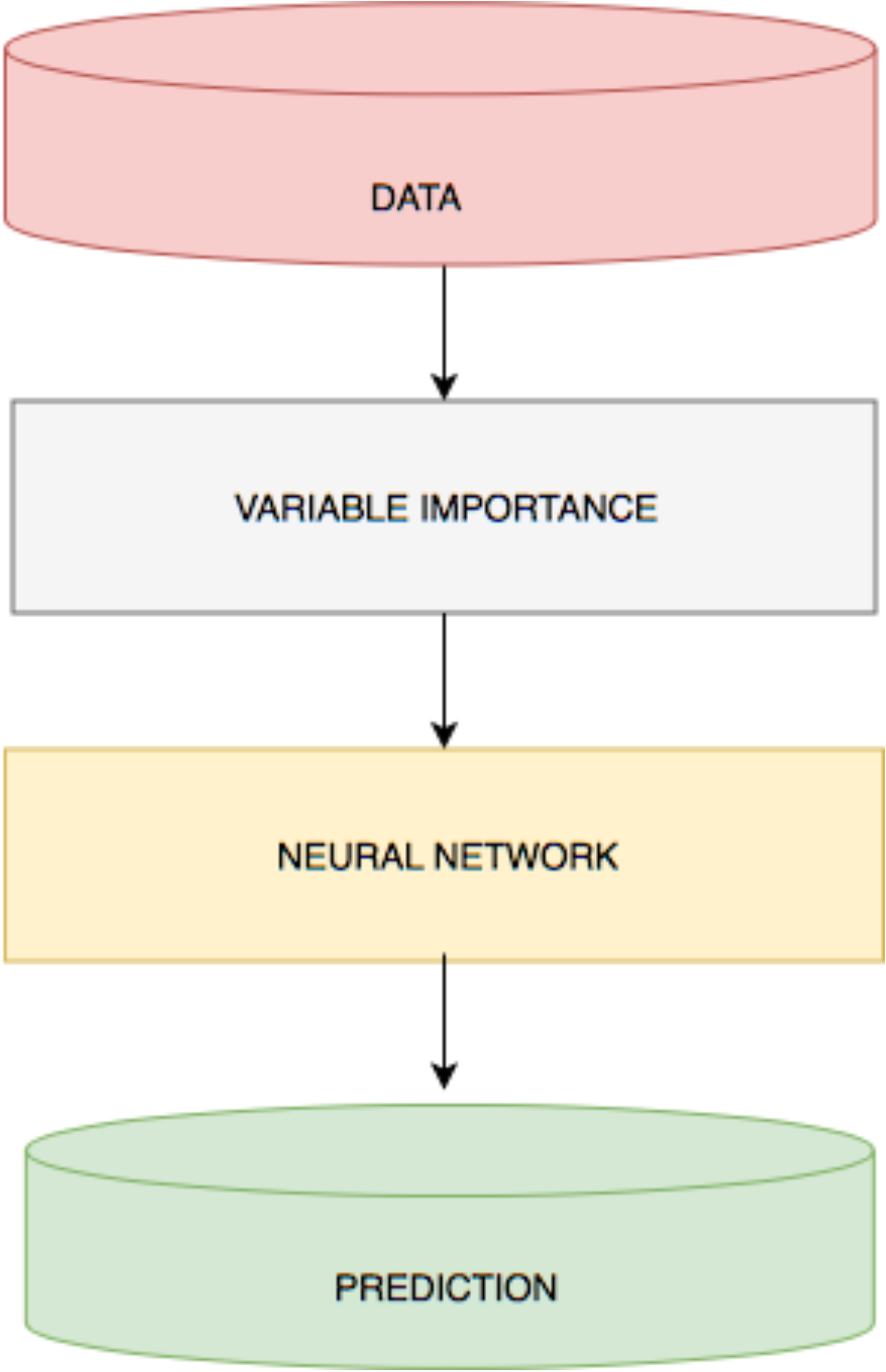

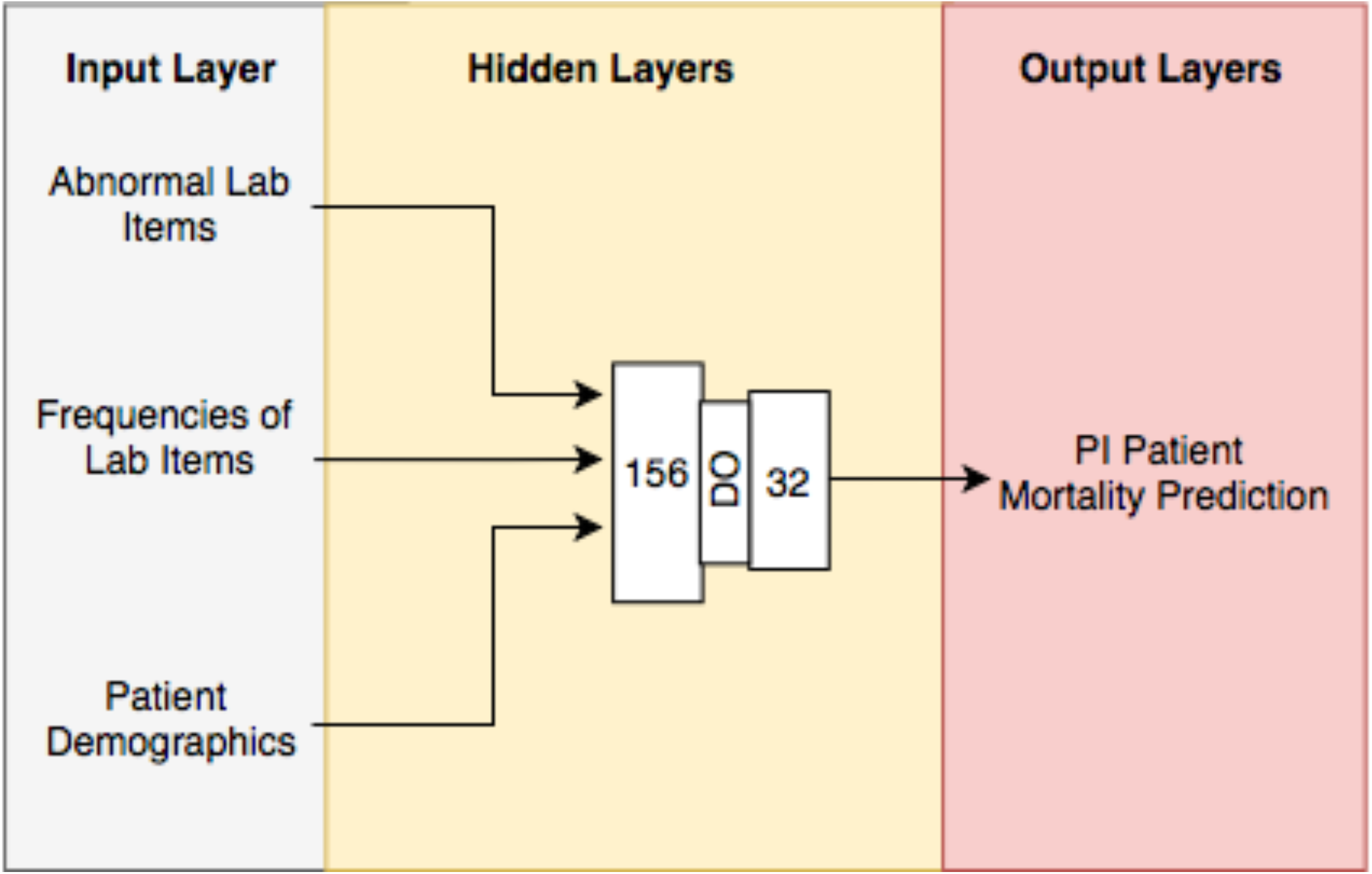

